# Q fever in Spain: epidemiology and demographic characteristics of hospitalized patients (2016–2023)

**DOI:** 10.64898/2026.07.09.26357673

**Authors:** Rafael Garcia-Carretero, Beatriz Valle-Borrego, Clara Peiro-Villalba, Maria-Dolores Martin-Rodrigo, Sara-Maria Quevedo-Soriano

**Affiliations:** Department of Internal Medicine, Hospital Universitario Severo Ochoa (Madrid, Spain); Department of Microbiology, Hospital Universitario Severo Ochoa (Madrid, Spain)

**Keywords:** *Coxiella burnetii*, mortality, hospitalization rate, Q fever

## Abstract

**Background:** Q fever, caused by Coxiella burnetii, is a zoonosis with significant public health implications. Spain has the highest number of cases in the European Union/European Economic Area, but the clinical and hospitalization burdens remain poorly characterized. This study described the epidemiology, demographic and clinical characteristics, and geographical distribution of hospitalized Q fever patients in Spain from 2016 to 2023.

**Methods:** We conducted a nationwide, retrospective study using the Spanish Minimum Basic Data Set for Hospitalization (MBDS-H). All hospital admissions with an ICD-10-CM code for Q fever (A78) between 2016 and 2023 were included. We analyzed demographic data, comorbidities, complications, length of stay, intensive care unit (ICU) admission, and mortality. We calculated hospitalization rates per 100,000 population. Temporal trends were assessed using Poisson regression.

**Results:** We identified 3,358 hospitalizations for Q fever, representing an overall hospitalization rate of 0.89 per 100,000 population. The median patient age was 56 years (interquartile range [IQR] 42–70), and the cohort was predominantly male (72%). The median hospital length of stay was 9 days (IQR 6–15), and 8.3% required ICU admission. The overall mortality rate was 2.4%. The most common complication was pneumonia (32%). Significant upward trends were observed over the study period for patient age, hypertension, and acute heart failure (p¡0.05). Geographical analysis revealed the highest hospitalization rates in the Canary Islands (2.33), La Rioja (2.16), and the Balearic Islands (1.93).

**Conclusion:** This study highlights the hospitalization burden due to Q fever in Spain. The risk of hospitalization increases with age and the presence of predisposing conditions. The marked regional heterogeneity and high frequency of complications such as pneumonia underscore the need for enhanced surveillance and a strengthened One Health approach to control this zoonosis.

## Introduction

Q fever is a bacterial infection caused by *Coxiella burnetii*, an intracellular pathogen. It is a zoonosis transmitted to humans via infected animals. While *C. burnetii* affects both domestic and wild animals, the primary reservoirs are domestic ruminants such as goats and sheep [1]. Consequently, Q fever is often considered an occupational disease, as it primarily affects farmers and workers in close contact with infected animals or animal products [2]. Humans may become infected through direct contact with the feces, urine, or other body fluids of these animals; however, the primary route of transmission is the inhalation of contaminated aerosols. C. burnetii is exceptionally resistant to adverse environmental conditions, allowing contaminated particles to travel long distances and cause outbreaks in populations far from the primary source of infection [1].

As an acute infectious disease, Q fever causes nonspecific signs and symptoms, typically characterized by the abrupt onset of fever, malaise, headache, and weakness; it is often included as a cause of fever of unknown origin. The disease can also present with severe complications, such as hepatitis, pneumonia, and cardiac or neurological involvement [1].

The prevalence of *C. burnetii* varies between countries and even within regions, depending on factors such as farming practices, animal populations, and occupational exposure [3]. Although the pathogen is found worldwide, the number of cases in the European Union/European Economic Area (EU/EEA) remained stable at 0.2 cases per 100,000 population between 2015 and 2019 [4, 5]. However, Spain reported the highest incidence of Q fever in the EU/EEA [4, 5], with 544 confirmed cases in 2024 (1.12 cases per 100,000 population) [6].

In Spain, Q fever is a notifiable disease. Coordination between epidemiological surveillance systems and veterinary services in livestock farming is essential for the control and prevention of the disease. While some recent studies describe the epidemiological landscape of Q fever in humans [6, 7, 8, 9], none have assessed the burden of hospitalization or characterized the clinical features and outcomes of *C. burnetii* infection.

The clinical presentation of *C. burnetii* infection depends on both the virulence of the pathogen’s strain and specific risk factors in the patient [1]. Studies on the clinical characteristics of hospitalized patients would help quantify the demand on hospital resources, identify trends over time, and characterize outcomes. To address this knowledge gap, we conducted a retrospective analysis of a national hospitalization database for the period 2016-2023.

The primary objective was to characterize the burden of hospitalization due to Q fever in Spain, including the number of admissions, patient demographics, and length of stay (LOS). Secondary objectives were to analyze trends in Q fever hospitalizations and to assess the geographical distribution across Spanish regions over an 8-year study period.

## Methods

### Study Design, Population, and Data Collection

A nationwide, retrospective study was conducted to describe the demographic and clinical characteristics of patients with Q fever in Spain between 2016 and 2023. Data were obtained from the Minimum Basic Data Set for Hospitalization (MBDS-H, or CMBD-H in Spanish). The MBDS-H is a mandatory registry based on discharge reports from all Spanish hospitals, both public and private. It is estimated that this registry covers nearly 95% of hospitals and collects about 97% of all discharge reports. As a standardized database of the Spanish National Health System, the MBDS-H provides data on the economic burden of hospital stays, as well as information regarding demographic characteristics, clinical outcomes, diagnoses, and diagnostic procedures. Since 2016, diagnoses in the MBDS-H have been coded using the 10th Clinical Modification of the International Classification of Diseases (ICD-10-CM). We identified 3,358 admissions in which Q fever was present. The resulting dataset included hospitalizations with anonymized patient records encoded using the ICD-10-CM code for Q fever (A78). While data from the MBDS-H are accessible from the Spanish Ministry of Health upon application, availability is typically delayed by 1 year. The Ministry provided a dataset with data through December 31, 2023.

### Selection of variables

Cases were identified by searching for the code A78 in any diagnostic position within the database. We selected demographic variables, such as sex, age, and place of residence, as well as dates of admission and discharge, clinical outcomes, and admission to the intensive care unit (ICU. Both complications and comorbidities were extracted, including hypertension, diabetes mellitus, acute heart failure, coronary disease, and malignancy. Regarding clinical complications, we included acute and subacute infective endocarditis (I33, I38, and I39), osteomyelitis (M86), acute hepatitis (B17 and B19.9), pneumonia (J18.0, J18.1, J18.8, and J18.9), and encephalitis, myelitis, and encephalomyelitis (G04).

### Statistical Analysis

We summarized the main characteristics of the population using descriptive statistics, including median and interquartile range (IQR), as well as absolute values and percentages. To provide a standardized measure of disease burden, we calculated absolute values and rates per 100,000 population for hospitalizations and mortality. Age-specific population data for these calculations were obtained from the Spanish National Institute of Statistics (Instituto Nacional de Estadística).

Poisson regression models were used to assess the significance of trends over time for Q fever-related admissions and mortality, with year treated as a continuous time variable. This approach allowed for the estimation of rate changes while avoiding the need for multiple comparison adjustments. For post hoc categorical comparisons (e.g., between age groups or comorbidities), we reported unadjusted p-values. A formal adjustment for multiple comparisons (e.g., Bonferroni) was not applied to the primary regression models, as the hypotheses focused on overall trends rather than individual pairwise tests. Consequently, the resulting p-values indicate whether a significant linear trend existed across the entire study period (2016-2023), rather than between individual years. A p-value ¡ 0.05 was considered statistically significant. All analyses were conducted using R version 4.2.1.

### Ethical Considerations

The study was conducted in compliance with the Declaration of Helsinki. Data were obtained through a formal request to the Spanish Ministry of Health. Patient records were provided anonymized and de-identified, ensuring confidentiality and privacy. Because no names or personal information were recorded, no additional patient consent was required.

## Results

During the 8-year study period from 2016 to 2023, 3,358 hospital admissions with a diagnosis of Q fever were identified in the Spanish MBDS-H database, representing an overall hospitalization rate (HR) of 0.89 cases per 100,000 population. The annual number of hospitalizations and the overall trend are depicted in Figure 1. There was an initial increase from 304 cases in 2016 to a peak of 491 in 2018. Although the number of annual admissions showed some fluctuation in subsequent years, it remained stable, with 402 cases recorded in 2023.

**Figure 1.**
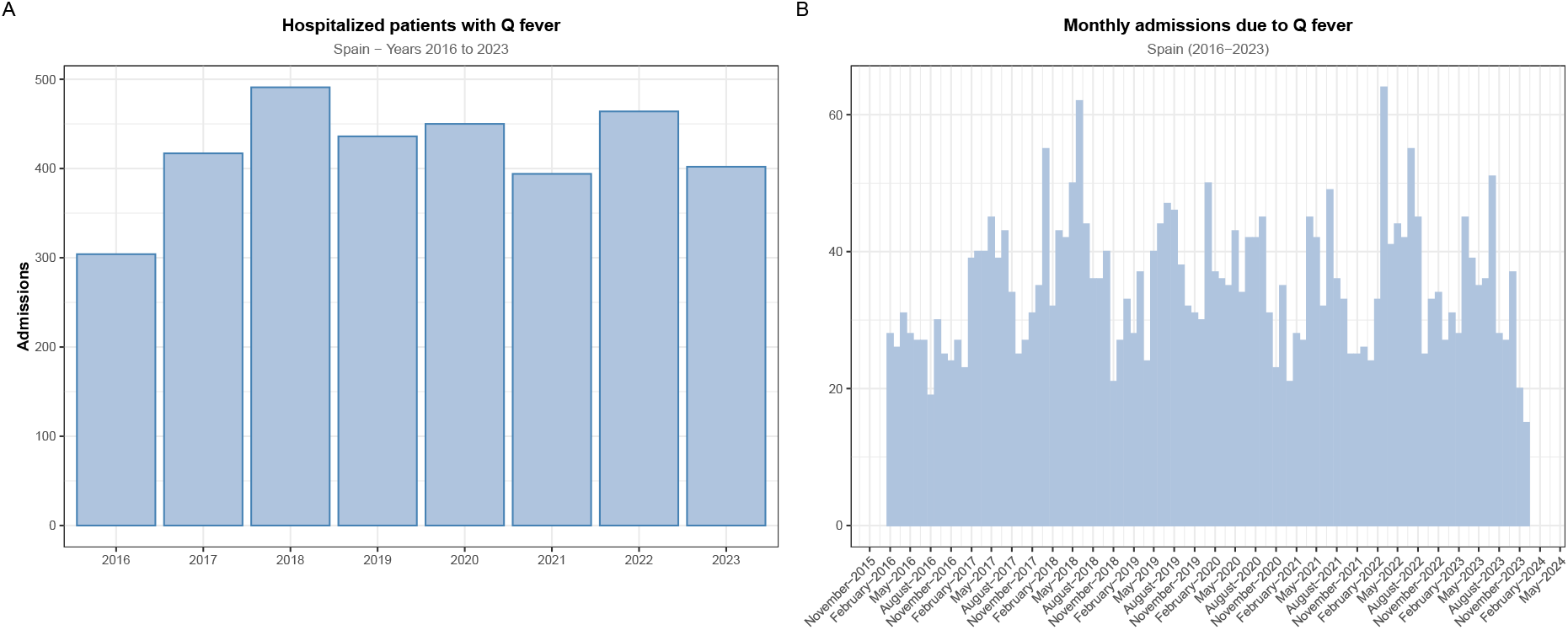
Trends in Q fever hospitalizations in Spain

### Demographic and clinical characteristics

The main characteristics of the hospitalized patients are summarized in Table . The median age was 56 years (IQR: 42–70), with a significant upward trend observed over the study period (p¡0.001). Most patients were men (2,429 patients; 72%), with no significant variation in sex distribution over the years (p=0.68).

The median LOS was 9 days (IQR: 6–15). In all, 278 patients (8.3%) required admission to the ICU, with a median ICU LOS of 5 days (IQR: 2–12). Neither ICU admissions nor ICU LOS demonstrated a significant trend over time. The overall in-hospital mortality rate was 2.4% (82 deaths), which remained stable over the study period.

### Comorbidities and complications

The most common comorbidities among hospitalized Q fever patients were hypertension (22%), type 2 diabetes mellitus (16%), and heart failure (9.8%). Over the study period, a significant increasing linear trend was observed for the prevalence of hypertension (p=0.043) and coronary disease (p=0.010). The Charlson Comorbidity Index (CCI) also significantly increased over the study period (p=0.042), with the proportion of patients in the highest comorbidity category (CCI *≥* 4) growing from 17% in 2016 to 22% in 2023.

Regarding acute complications, pneumonia was the most frequent, occurring in 1,060 patients (32%). Acute hepatitis was documented in 2.7% of cases, and infective endocarditis was recorded in 130 patients (3.9%). Neurological (encephalitis/encephalomyelitis) and bone (osteomyelitis) complications were rare, occurring in less than 0.5% of the cohort. Co-infection with Severe Acute Respiratory Syndrome Coronavirus 2 was present in 83 patients (2.5%), with a highly significant trend (p¡0.001), peaking at 8.0% in 2022.

### Geographical distribution

The geographical distribution of Q fever hospitalizations was heterogeneous (Table 1, Figure 2). The highest cumulative hospitalization rates were observed in the Canary Islands (2.33 per 100,000), La Rioja (2.16), and the Balearic Islands (1.93). In contrast, regions such as Catalonia (0.37), Cantabria (0.90), and Castilla - La Mancha (0.65) reported the lowest rates. Several regions, including Extremadura, Aragón, and Asturias, showed year-to-year variability in their incidence rates.

**Table 1.** Annual hospitalization rates for Q fever per 100,000 population by Spanish region, 2016–2023.

| Region | 2016 | 2017 | 2018 | 2019 | 2020 | 2021 | 2022 | 2023 | Total |
| --- | --- | --- | --- | --- | --- | --- | --- | --- | --- |
| Andalucía | 49 (0.58) | 91 (1.09) | 76 (0.91) | 81 (0.96) | 60 (0.71) | 58 (0.68) | 72 (0.85) | 81 (0.96) | 568 (0.84) |
| Aragón | 10 (0.76) | 16 (1.22) | 32 (2.45) | 14 (1.06) | 34 (2.56) | 19 (1.43) | 11 (0.83) | 11 (0.83) | 147 (1.39) |
| Asturias | 7 (0.67) | 11 (1.06) | 25 (2.43) | 12 (1.17) | 13 (1.28) | 3 (0.3) | 24 (2.37) | 18 (1.78) | 113 (1.38) |
| Illes Balears | 5 (0.45) | 23 (2.06) | 43 (3.81) | 21 (1.83) | 26 (2.22) | 20 (1.71) | 20 (1.71) | 19 (1.62) | 177 (1.93) |
| Canarias | 45 (2.14) | 45 (2.13) | 51 (2.4) | 55 (2.55) | 58 (2.67) | 58 (2.67) | 51 (2.35) | 38 (1.75) | 401 (2.33) |
| Cantabria | 0 (0) | 6 (1.03) | 8 (1.38) | 8 (1.38) | 5 (0.86) | 7 (1.2) | 7 (1.2) | 1 (0.17) | 42 (0.9) |
| Castilla y León | 3 (0.12) | 1 (0.04) | 33 (1.37) | 28 (1.17) | 18 (0.75) | 22 (0.92) | 34 (1.43) | 16 (0.67) | 155 (0.81) |
| Castilla – La Mancha | 7 (0.34) | 13 (0.64) | 12 (0.59) | 13 (0.64) | 14 (0.68) | 17 (0.83) | 19 (0.93) | 11 (0.54) | 106 (0.65) |
| Catalunya | 14 (0.19) | 23 (0.3) | 33 (0.43) | 31 (0.4) | 21 (0.27) | 34 (0.44) | 41 (0.53) | 29 (0.37) | 226 (0.37) |
| Comunitat Valenciana | 37 (0.75) | 33 (0.67) | 26 (0.52) | 24 (0.48) | 29 (0.57) | 21 (0.42) | 42 (0.83) | 44 (0.87) | 256 (0.64) |
| Extremadura | 11 (1.01) | 8 (0.74) | 12 (1.12) | 14 (1.31) | 42 (3.95) | 28 (2.64) | 23 (2.17) | 22 (2.08) | 160 (1.87) |
| Galicia | 21 (0.77) | 17 (0.63) | 19 (0.7) | 15 (0.56) | 26 (0.96) | 17 (0.63) | 24 (0.89) | 15 (0.56) | 154 (0.71) |
| Madrid | 48 (0.74) | 58 (0.89) | 58 (0.88) | 48 (0.72) | 44 (0.65) | 36 (0.53) | 37 (0.55) | 35 (0.52) | 364 (0.68) |
| Murcia | 16 (1.09) | 20 (1.36) | 18 (1.22) | 19 (1.27) | 15 (0.99) | 16 (1.05) | 13 (0.86) | 21 (1.38) | 138 (1.15) |
| Nafarroa | 9 (1.4) | 5 (0.78) | 8 (1.24) | 8 (1.22) | 6 (0.91) | 6 (0.91) | 8 (1.21) | 8 (1.21) | 58 (1.11) |
| Euskadi | 15 (0.69) | 30 (1.37) | 26 (1.18) | 32 (1.45) | 36 (1.62) | 30 (1.36) | 37 (1.67) | 27 (1.22) | 233 (1.32) |
| La Rioja | 6 (1.9) | 14 (4.44) | 10 (3.17) | 13 (4.1) | 3 (0.94) | 2 (0.63) | 1 (0.31) | 6 (1.88) | 55 (2.16) |
| Ceuta | 1 (1.18) | 2 (2.35) | 0 (0) | 0 (0) | 0 (0) | 0 (0) | 0 (0) | 0 (0) | 3 (0.45) |
| Melilla | 0 (0) | 1 (1.16) | 1 (1.16) | 0 (0) | 0 (0) | 0 (0) | 0 (0) | 0 (0) | 2 (0.29) |
| Total | 304 (0.65) | 417 (0.9) | 491 (1.05) | 436 (0.93) | 450 (0.95) | 394 (0.83) | 464 (0.98) | 402 (0.85) | 3358 (0.89) |

Data are expressed as number of cases (hospitalization rate per 100,000 population)

**Figure 2.**
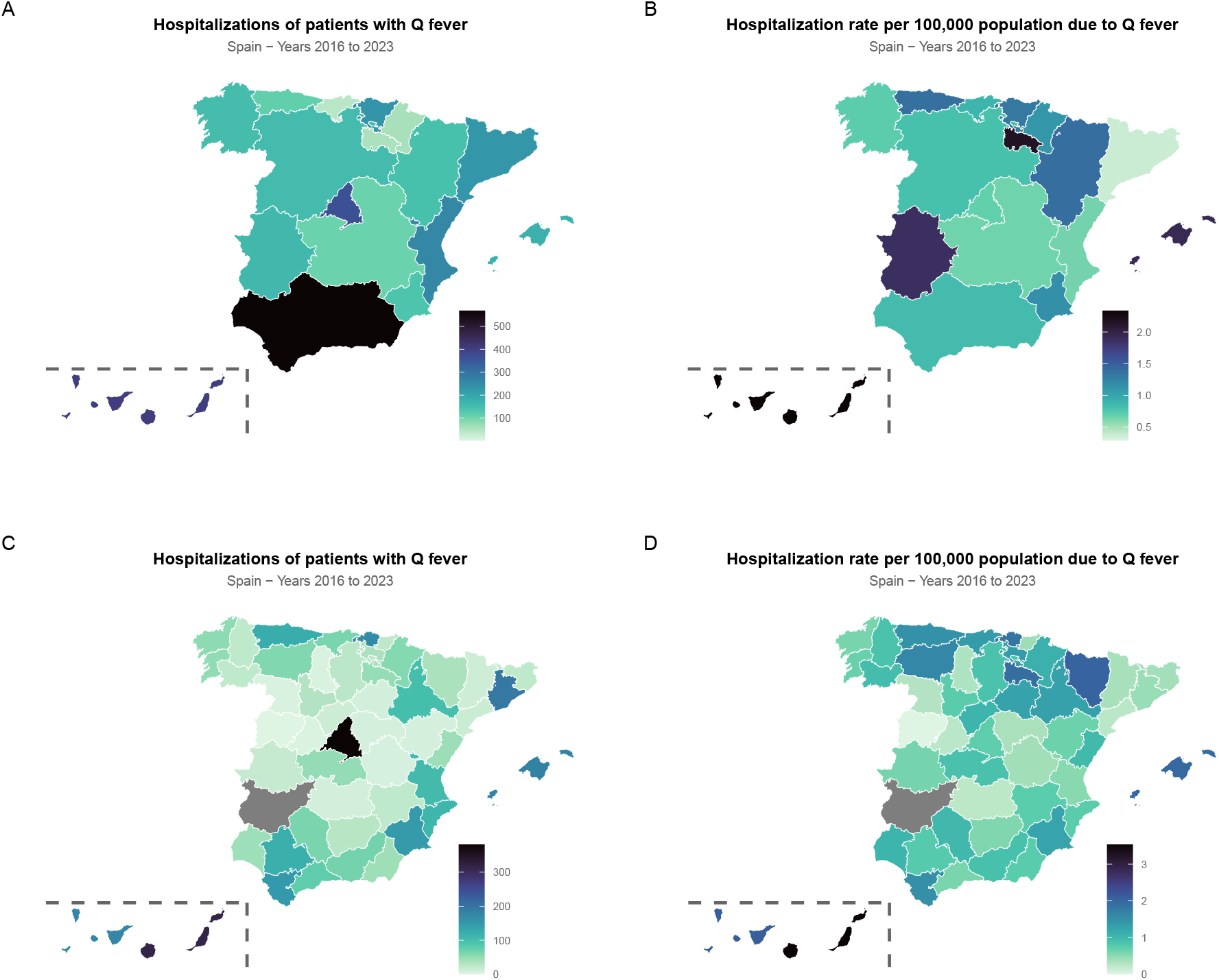
Regional and provincial distribution of Q fever hospitalizations in Spain, 2016–2023: (A) absolute number of cases by region; (B) hospitalization rate by region; (C) absolute number of cases by province; and (D) hospitalization rate by province.

### Age-specific hospitalization rates

The relationship between age and disease burden is shown in Table 2 and Figure 3. The number of cases and the hospitalization rate increased steadily with age. The lowest rates were observed in pediatric age groups (e.g., 0.16 per 100,000 in children under 4 years). Rates climbed consistently starting from the 25–29 years age group (0.67) and peaked among those aged 80–84 years (1.77 per 100,000), followed by a slight reduction in those aged 85 years and older (1.46).

**Table 2.**
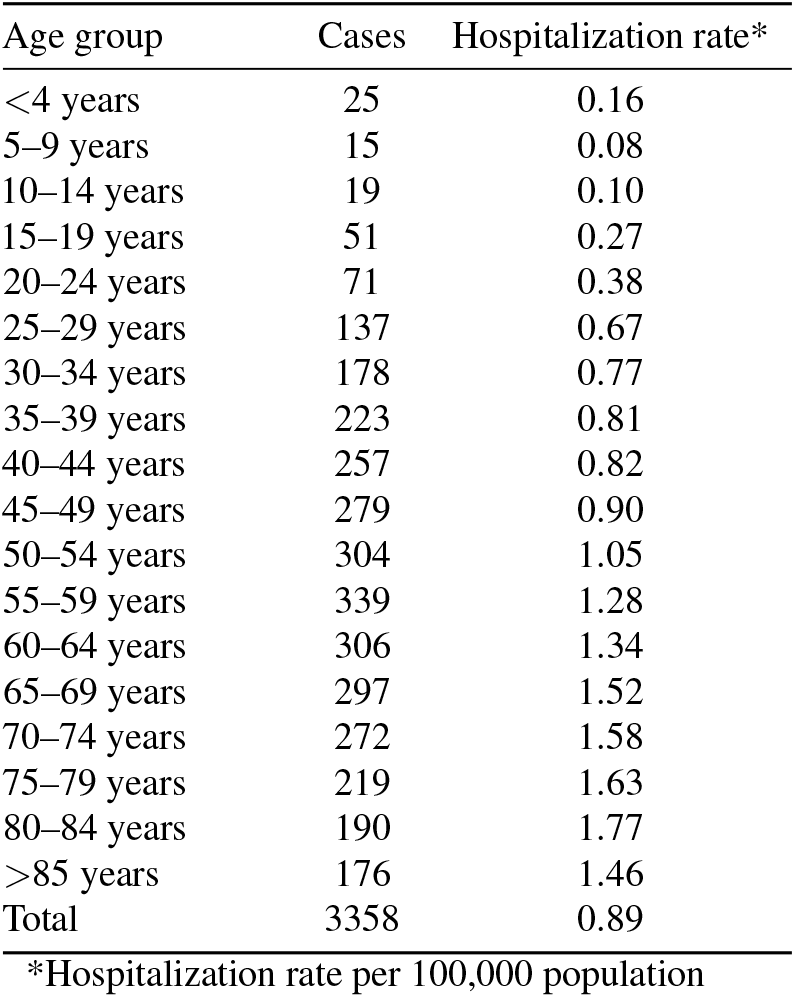
Q fever cases and age-adjusted incidence (measured as hospitalization rate per 100,000 population) in Spain, 2016–2023.

| Age group | Cases | Hospitalization rate* |
| --- | --- | --- |
| <4 years | 25 | 0.16 |
| 5–9 years | 15 | 0.08 |
| 10–14 years | 19 | 0.10 |
| 15–19 years | 51 | 0.27 |
| 20–24 years | 71 | 0.38 |
| 25–29 years | 137 | 0.67 |
| 30–34 years | 178 | 0.77 |
| 35–39 years | 223 | 0.81 |
| 40–44 years | 257 | 0.82 |
| 45–49 years | 279 | 0.90 |
| 50–54 years | 304 | 1.05 |
| 55–59 years | 339 | 1.28 |
| 60–64 years | 306 | 1.34 |
| 65–69 years | 297 | 1.52 |
| 70–74 years | 272 | 1.58 |
| 75–79 years | 219 | 1.63 |
| 80–84 years | 190 | 1.77 |
| >85 years | 176 | 1.46 |
| Total | 3358 | 0.89 |

\*Hospitalization rate per 100,000 population

**Figure 3.**
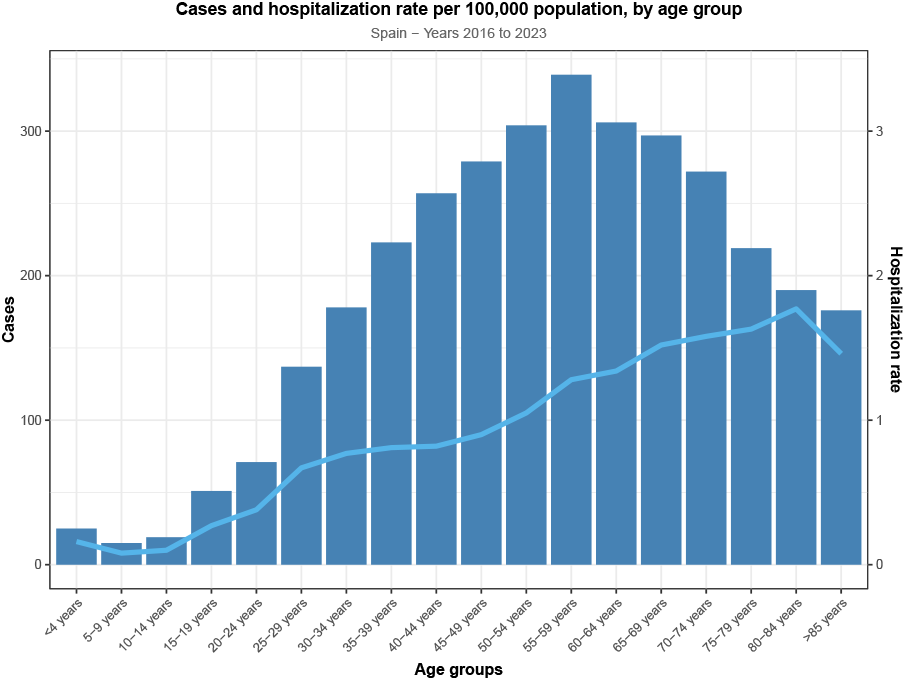
Cases of Q fever (bars) and hospitali zation rate (blue line) per 100,000 population in Spain, 2016–2023.

## Discussion

We conducted a nationwide, retrospective study to provide a comprehensive analysis of the incidence, hospitalizations, and clinical characteristics of confirmed cases of Q fever in Spain over an 8-year period. We identified 3,358 patients during the observation period, representing an HR of 0.89 per 100,000 population. In 2016, the HR was 0.65, compared to 0.53 prior to 2016 [8]. Following 2016, both the number of confirmed cases and the HR remained stable across the study period. This initial increase may be attributed to the classification of Q fever as a notifiable disease in Spain beginning in 2015.

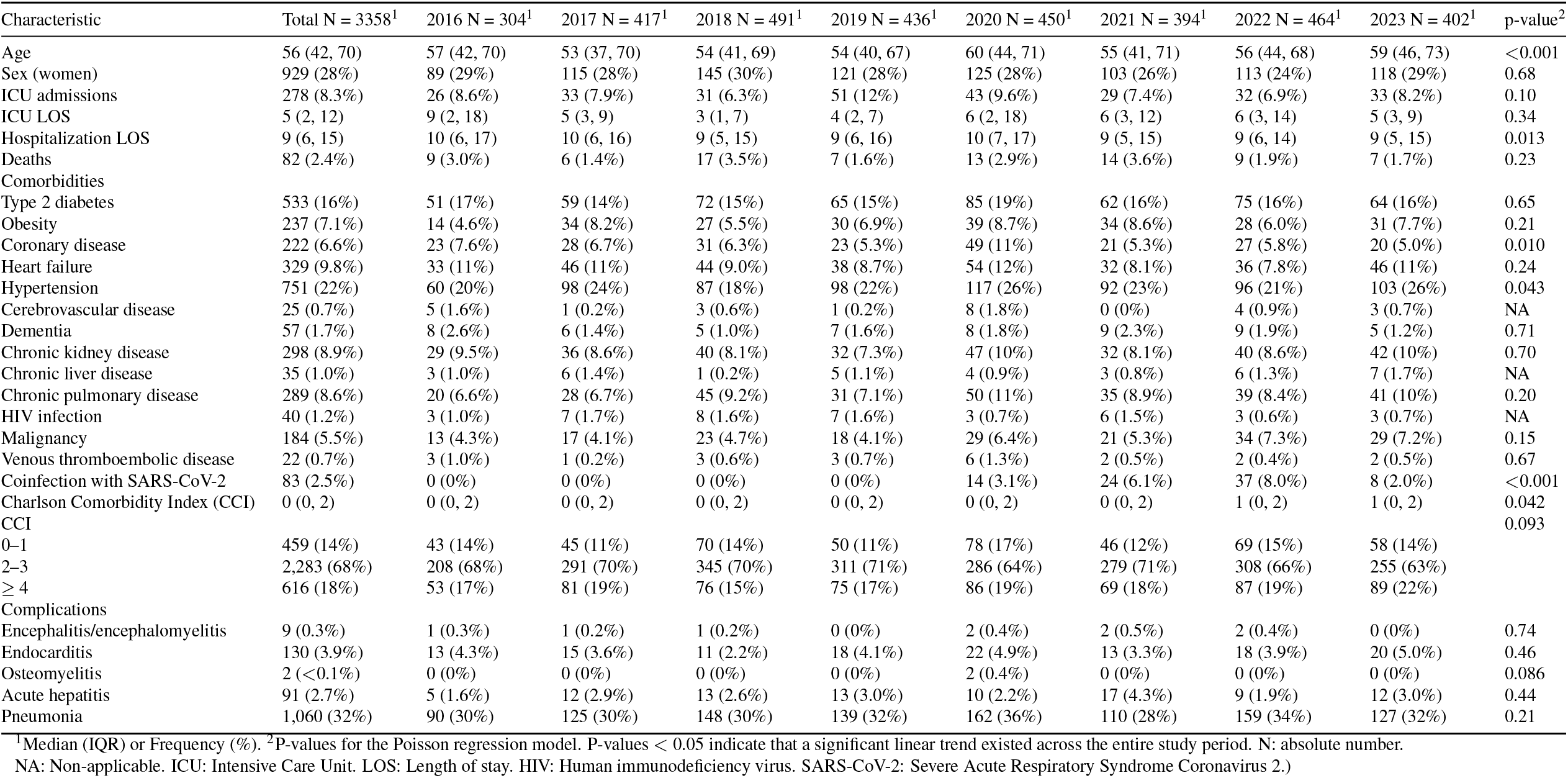

It is worth noting the relationship between Spain’s two primary registries, RENAVE (the Spanish National Epidemiological Surveillance Network) [6] and the MBDS-H. The former aggregates demographic data on confirmed, notified cases regardless of hospitalization status, whereas the MBDS-H contains demographic and clinical information specifically for hospitalized patients. A recent study that compared these registries between 2016 and 2022 [8] found that RENAVE was less comprehensive than the MBDS-H, revealing underreporting (2,429 cases vs. 2,991 cases). This discrepancy may stem from the progressive adherence of different regions to the notification system. Furthermore, while some regions have not fully integrated into the system, it is also possible that mild or asymptomatic cases were not registered by RENAVE but were subsequently captured in the MBDS-H via hospital discharge reports [8].

While the trend remained stable from 2017 onward, a slight decrease in cases was recorded in 2021. This is likely explained by the COVID-19 pandemic and subsequent underdiagnosis or underreporting due to the limited availability of healthcare services [6]. Consistent with previous studies [1, 5, 6], we identified a seasonal trend with a peak in confirmed cases from March to July, which is likely associated with the livestock breeding season [10, 11]. Additionally, some authors suggest that environmental factors during these months create favorable conditions, specifically regarding humidity and temperature, for the dispersal of *C. burnetii* pseudo-spores [8, 9].

Nevertheless, Spain has reported the highest number of notified cases in Europe since 2017 [4]. According to the 2019 Annual Epidemiological Report from the European Centre for Disease Prevention and Control (ECDC), Spain accounted for more than one-third of confirmed cases, with 332 individuals [4], followed by Germany (155) and France (148), as depicted in Figure 4. In the same year, the notification rate in Spain was 0.7 per 100,000 population, compared to 0.2 in the EU/EEA. This disparity may be attributed to the fact that Q fever is endemic in Spain and that the national notification system has improved since 2015 [4].

**Figure 4.**
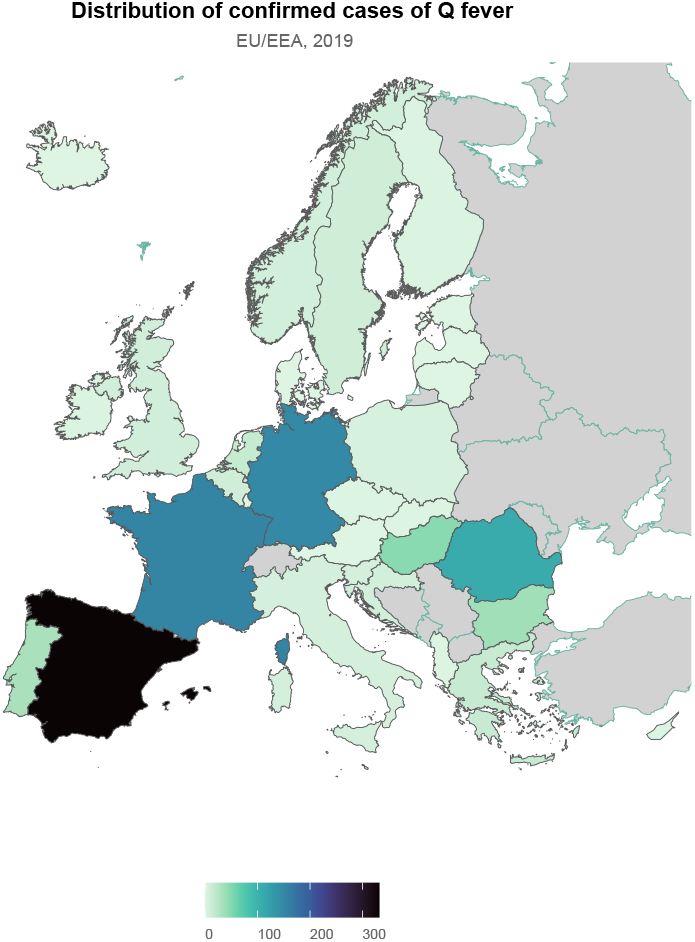
Distribution of confirmed cases of Q fever in countries in the EU/EEA (data from 2019).

We found a predominance of middle-aged males (72% men vs. 28% women; median age: 56 years). This highlights the occupational nature of *C. burnetii* infection, as livestock activity has traditionally been associated with men [12]. Furthermore, our findings suggest that age and specific comorbidities are associated with hospitalizations. Although the absolute number of cases peaked at 55–59 years, the HR steadily increased up to until the 80–84 age group, reaching a peak of 1.77 per 100,000 population. The most prevalent comorbidities were hypertension (22%), type 2 diabetes mellitus (16%), and heart failure (9.8%), with rates increasing significantly over the study period. This trend highlights the association between advancing age and these comorbidities. Likewise, the CCI showed significantly increased over the study period, which may reflect the higher clinical severity in older adults that leads to hospitalization [8].

Regarding complications, pneumonia was the most frequent, occurring in 32% of cases. Endocarditis and hepatitis (3.9% and 2.7%, respectively) were also prevalent among hospitalized patients. These findings fall within the reported ranges for acute and chronic Q fever complications [1], emphasizing the critical importance of early identification and monitoring of complications in high-risk patients [13]. Finally, the low mortality rate (2.4%) suggests that appropriate healthcare and early screening for complications may improve Q fever outcomes.

The heterogeneity in the HR across different regions in Spain is noteworthy. We found consistently high rates in the Canary Islands (2.33), La Rioja (2.16), the Balearic Islands (1.93), and Extremadura (1.87). Except for La Rioja, a decreasing trend was observed in these high-rate regions over the study period. This finding has also been reported in other Spanish studies [6, 7]. More interestingly, when the data were broken down to a provincial level, certain provinces had the highest number of cases: Las Palmas (315), Barcelona (194), Santa Cruz de Tenerife (172), Vizcaya (168), and Cádiz (150). This may reflect endemic factors related to specific environmental, zoonotic, and livestock characteristics, as pointed out by previous studies [7, 9, 13]. This heterogeneity highlights the need for targeted, region-specific public health interventions. Enhanced surveillance and coordination between human and veterinary health services (a *One Health* approach) in these high-incidence regions should be the focus for outbreak prevention and control.

### Limitations

This study had several limitations inherent to its design and data source. Our results relied on an administrative database that depended on diagnostic coding accuracy; therefore, mis-coding or underreporting could affect our estimates. Consequently, the findings may reflect potential coding inaccuracies or omissions. The MBDS-H only captures the most severe cases requiring hospitalization and should not be considered a gold standard or a substitute for the RENAVE registry, as it may overlook mild or asymptomatic Q fever infections that do not require admission. However, considering the nationwide nature of the study, the large sample size, and the consistent trends observed across multiple years, we believe that our findings are robust.

We are also aware that the database lacks detailed clinical, microbiological, and exposure data. This prevents us from analyzing specific risk factors, treatment responses, or outcomes related to chronic Q fever beyond the initial hospitalization, which would provide additional insight into the factors driving hospitalization and mortality trends. Because the database is built upon discharge reports, we cannot account for changes in coding practices, clinical guidelines, diagnostic criteria, or data collection methods over time. Finally, the impact of COVID-19 on hospitalizations, diagnostic criteria, clinical management, and outcomes during 2020–2023 may represent a confounding factor that could had led to the misinterpretation of trends due to pandemic-related effects.

## Conclusion

To the best of our knowledge, this is the first study to summarize the demographic and clinical characteristics and the burden of Q fever hospitalizations in Spain. Our study quantifies a significant burden of hospitalizations, characterized by distinct demographic, clinical, and geographical patterns. The increasing age and comorbidity burden of hospitalized patients, the high rate of acute and chronic complications, and the marked regional disparities highlight the need for continued and enhanced surveillance in high-incidence areas. Finally, our findings emphasize the need for strategies to control animal reservoirs and reduce the impact of this zoonosis on human health in Spain.

## Data Availability

The datasets generated during and/or analysed during the current study are not publicly available because a contract signed with the Spanish National Health System, which provided the data set, prohibits the authors from providing their data to any other researcher. Furthermore, the authors must destroy the database upon the conclusion of their investigation. The database cannot be uploaded to any public repository. However, they are are available on reasonable request at https://www.sanidad.gob.es/estadEstudios/estadisticas/sisInfSanSNS/aplicacionesConsulta/home.htm.

## Declarations

### Authors’ contributions

Dr. Garcia-Carretero conceived and designed the study, wrote the first draft of the manuscript, and preprocessed and analyzed the data. Dr. Valle-Borrego, Dr. Peiro-Villalba and Dr. Martin-Rodrigo made substantial contributions to the interpretation of the results, critically reviewed the first draft of the manuscript, made valuable suggestions, and contributed to the visualization of the data. Drs. Quevedo-Soriano supervised the project and critically reviewed and edited the final draft of the manuscript. All authors read and approved the final manuscript.

### Availability of data and materials

A contract signed with the Spanish National Health System, which provided the dataset, prohibits the authors from providing their data to any other researcher. Furthermore, the authors must destroy the database upon the conclusion of their investigation. The database cannot be uploaded to any public repository.

### Ethics approval and consent to participate

No identifying information was included in the manuscript. Because the authors used historical data, informed consent was not necessary and no ethical approval was required. All procedures involving human participants were conducted in accordance with the ethical standards of the responsible institutional and/or national research committee and with the 1964 Helsinki Declaration and its later amendments or comparable ethical standards.

### Consent for publication

Not applicable.

### Declaration of generative AI and AI-assisted technologies in the manuscript preparation process

During the preparation of this work the authors did not use AI-assisted technologies. The authors take full responsibility for the content of the published article.

